# The Fecal Microbiome in Quiescent Crohn’s Disease with Persistent Gastrointestinal Symptoms Show Enrichment of Oral Microbes But Depletion of Butyrate and Indole Producers

**DOI:** 10.1101/2023.05.16.23290065

**Authors:** Jonathan Golob, Krishna Rao, Jeffrey Berinstein, William Chey, Chung Owyang, Nobuhiko Kamada, Peter Higgins, Vincent Young, Shrinivas Bishu, Allen Lee

## Abstract

**Background and Aims:** Even in the absence of inflammation, persistent symptoms in Crohn’s disease (CD) are prevalent and negatively impact quality of life. We aimed to determine whether quiescent CD patients with persistent symptoms (**qCD+symptoms**) have changes in microbial structure and functional potential compared to those without symptoms (**qCD-symptoms**).

**Methods:** We performed a prospective multi-center observational study nested within the SPARC IBD study. CD patients were included if they had evidence of quiescent disease as defined by fecal calprotectin level < 150 mcg/g. Persistent symptoms were defined by the CD-PRO2 questionnaire. Active CD (**aCD**), diarrhea-predominant irritable bowel syndrome (**IBS-D**), and healthy controls (**HC**) were included as controls. Stool samples underwent whole genome shotgun metagenomic sequencing.

**Results:** A total of 424 patients were analyzed, including 39 qCD+symptoms, 274 qCD-symptoms, 21 aCD, 40 IBS-D, and 50 HC. Patients with qCD+symptoms had a less diverse microbiome, including significant reductions in Shannon diversity (*P*<.001) and significant differences in microbial community structure (*P*<.0001), compared with qCD-symptoms, IBS-D, and HC. Further, patients with qCD+symptoms showed significant enrichment of bacterial species that are normal inhabitants of the oral microbiome, including *Klebsiella pneumoniae* (q=.003) as well as depletion of important butyrate and indole producers, such as *Eubacterium rectale* (q=.001), *Lachnospiraceae spp*. (q<.0001), and *Faecalibacterium prausnitzii* (q<.0001), compared with qCD-symptoms. Finally, qCD+symptoms showed significant reductions in bacterial *tnaA* genes, which mediate tryptophan metabolism, as well as significant *tnaA* allelic variation, compared with qCD-symptoms.

**Conclusion:** The microbiome in patients with qCD+symptoms show significant changes in diversity, community profile, and composition compared with qCD-symptoms. Future studies will focus on the functional significance of these changes.

**What You Need to Know:** *Background:* Persistent symptoms in quiescent Crohn’s disease (CD) are prevalent and lead to worse outcomes. While changes in the microbial community have been implicated, the mechanisms by which altered microbiota may lead to qCD+symptoms remain unclear.

*Findings:* Quiescent CD patients with persistent symptoms demonstrated significant differences in microbial diversity and composition compared to those without persistent symptoms. Specifically, quiescent CD patients with persistent symptoms were enriched in bacterial species that are normal inhabitants of the oral microbiome but depleted in important butyrate and indole producers compared to those without persistent symptoms.

*Implications for Patient Care:* Alterations in the gut microbiome may be a potential mediator of persistent symptoms in quiescent CD. Future studies will determine whether targeting these microbial changes may improve symptoms in quiescent CD.

## Introduction

Although modern therapies have improved care in inflammatory bowel disease (IBD), improving inflammation does not improve quality of life in many IBD patients.^1^ Even in the absence of active inflammation, persistent gastrointestinal symptoms are reported in up to 46% of IBD patients, particularly with Crohn’s disease (CD).^2^ Quiescent CD with persistent GI symptoms (**qCD+symptoms**) result in significant deterioration in quality of life similar to active inflammation.^3^ Furthermore, qCD+symptoms are independent risk factors for future opioid use,^4^ associated with higher mortality,^5^ and among the top factors associated with high costs of IBD care.^6^ Despite this, mechanisms underpinning this condition are poorly understood and evidence-based therapies are limited.

Prior studies have demonstrated disrupted epithelial barrier integrity in qCD+symptoms.^7,8^ Although initially attributed to subclinical inflammation, the prevalence of persistent symptoms is similar in quiescent IBD patients with and without deep remission.^9^ More recently, altered microbial communities have been described in quiescent IBD with persistent diarrhea,^10^ psychological distress,^11^ and fatigue.^12^ While these studies raise the possibility of disturbed brain-gut-microbiome axes, the mechanisms by which an altered microbiome may mediate qCD+symptoms are still largely unexplored. Intriguingly, quiescent IBD patients with fatigue were noted to have decreased serum tryptophan levels.^12^ Tryptophan is a key regulator of host-microbial interactions and influences intestinal barrier function, visceral sensation, and brain-gut interactions.^13^ Thus, we hypothesized that qCD+symptoms would exhibit altered intestinal microbial communities, particularly with microbial genes important in tryptophan metabolism.

In this study, we aimed to identify structural and functional alterations in the fecal microbiota and determine whether changes in microbial genes important in tryptophan pathways were seen in qCD+symptoms compared to quiescent CD without symptoms (**qCD-symptoms**).

## Materials and Methods

### Patient Cohort

We performed a prospective multi-center observational study nested within the Study of a Prospective Adult Research Cohort with Inflammatory Bowel Disease (SPARC IBD), which is a prospective longitudinal cohort study of adult IBD patients across the US utilizing standardized collection methods and sample processing techniques.^14^ Patients were included if they had an established diagnosis of CD, ≥18 years of age, and had quiescent disease as defined by fecal calprotectin (FCP) <150 mcg/g, which correlates well with endoscopic healing and suggested as treatment targets by consensus guidelines.^15^ Persistent symptoms were defined as mean abdominal pain score ≥2 and daily liquid stool frequency ≥4 using the CD-PRO2 score, which were suggested as clinically meaningful targets by consensus guidelines.^15,16^ FCP levels had to be drawn within +/-4 weeks of completing CD-PRO2 scores. Exclusion criteria included history of total colectomy or presence of ileostomy or colostomy.

As controls, we included patients from SPARC IBD with active CD (**aCD**) defined by FCP >150 mcg/g as well as non-IBD patients, including diarrhea-predominant irritable bowel syndrome (**IBS-D**) and healthy controls (**HC**). IBS-D were required to meet Rome IV criteria while HC had absence of any active/chronic medical condition. All patients provided written informed consent prior to enrollment. The institutional review board at the University of Michigan gave ethical approval for this work. All authors had access to the study data and reviewed and approved the final manuscript.

### Metagenomic Sequencing

All patients had stool samples available which underwent whole genome shotgun (WGS) metagenomic sequencing. Raw WGS reads were then assembled *de novo* and underwent taxonomic and functional annotation using *geneshot*.^17^ Please see supplemental methods for full details.

### Statistical Analysis

Continuous variables were compared using t-tests or one-way analysis of variance (ANOVA) and Tukey’s test for post-hoc analyses. Chi-square or Fisher’s exact test was utilized to compare categorical variables. A two-tailed *P*<.05 was considered significant. Adjustment for multiple comparisons was performed using the Benjamini-Hochberg method (target false discovery rate q<.05). All analyses were performed using R (version 4.2.1).

Diversity metrics were calculated using the R package *vegan*, including Shannon index as a measure of microbial diversity within-samples (α-diversity) and Jaccard distance for between-sample differences (β-diversity). Ordination analyses were performed using uniform manifold approximation and projection (UMAP) on Jaccard distance matrices at the species level. Differences in β-diversity metrics between groups were compared using permutational analysis of variance (PERMANOVA) implemented by the adonis function in *vegan* using 10,000 permutations. Differential abundance of microbial species was performed using the R package *ANCOM-BC*.^18^ Functional profiling of the microbial community was performed using HUMAnN3 based on the KEGG Orthology (KO) database.^19^ KO count contingency tables were constructed by presence/absence of genes and compared by Fisher’s exact test.

Finally, we performed random forest classification using the R package *tidymodels* to determine whether microbial species could predict qCD+symptoms vs. qCD-symptoms. Data were randomly partitioned with 75% of the data used for model training and 25% held out for testing and validation. Five-fold cross-validation was employed to estimate model accuracy and to tune hyperparameters. The prediction accuracy of the model was evaluated on the independent test set by calculating the area under the receiver operator characteristic curve (AUC). Variable importance scores were extracted to determine species most important in the model.

## Results

### Baseline Demographics

A total of 334 CD patients, including 39 qCD+symptoms, 274 qCD-symptoms, and 21 aCD, as well as 40 IBS-D and 50 HC had metagenomic data available for analysis (**Figure 1A, Table 1**).

**Figure 1.**
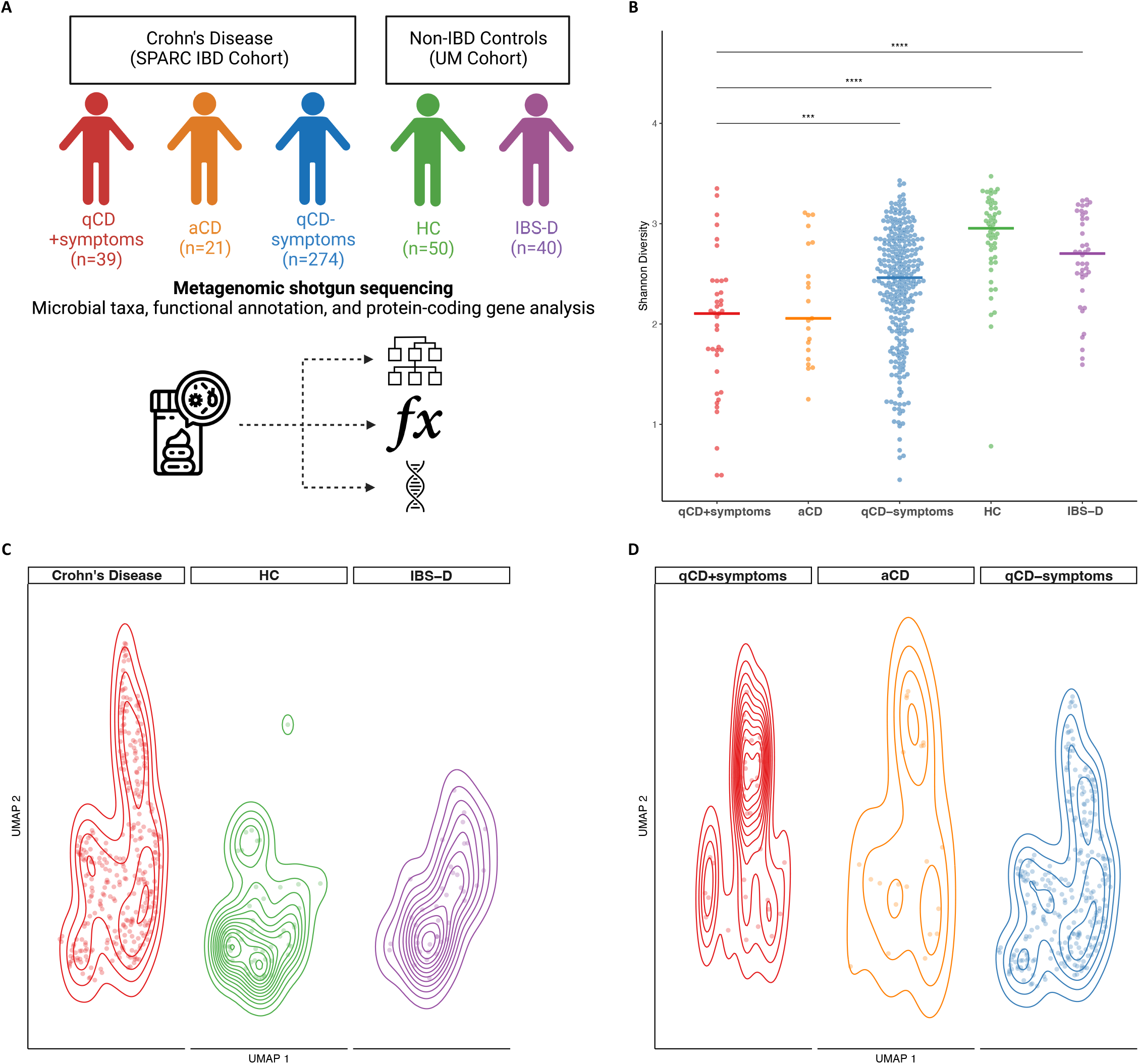
The microbiome in qCD+symptoms is similar to aCD but distinct from qCD-symptoms. (**A**) A total of 424 patients underwent WGS metagenomic analysis, including 39 with quiescent CD with persistent GI symptoms (qCD+symptoms, *red*), 21 with active CD (aCD, *orange*), and 274 with qCD without symptoms (qCD-symptoms, *blue*), 40 with diarrhea-predominant irritable bowel syndrome (IBS-D, *green*) and 50 healthy controls (HC, *purple*). (**B**) Shannon diversity was significantly reduced in qCD+symptoms compared with qCD-symptoms, IBS-D, and HC. Horizontal line represents median value for each group. *** adjusted *P*<.001; **** adjusted *P*<.0001. (**C**) There were significant differences in Jaccard distance at the species level in CD (*red*), HC (*green*), and IBS-D (*purple*) (*P*<.00001 by PERMANOVA). Each dot represents an individual subject ordinated by uniform manifold approximation and projection (UMAP) overlaid with 2D-contour plot. (**D**) When examining only CD subgroups, differences in microbial community structure were largely driven by qCD+symptoms (*red*), which was significantly different compared with qCD-symptoms (*blue*, adjusted P<.0001), but not compared with aCD (*orange*, adjusted *P*=.37).

### qCD+symptoms showed significant changes in microbial diversity

There were significant differences in Shannon index between the groups (**Figure 1B**, *P*<.0001) with qCD+symptoms demonstrating significantly decreased Shannon diversity compared with qCD-symptoms, IBS-D, and healthy controls (adjusted *P*<.001; *P*<.0001; and *P*<.0001, respectively).

We also found significant differences in microbial community between CD, IBS-D, and healthy controls (**Figure 1C**, *P*<.00001 by PERMANOVA). When focusing on CD subgroups, the microbial community structure in qCD+symptoms was significantly different compared with qCD-symptoms (**Figure 1D**, adjusted *P*<.0001) but not with aCD (adjusted *P*=.37).

### Changes in microbial diversity in qCD+symptoms were independent from inflammation

We next sought to determine whether our threshold for quiescent inflammation may have influenced our findings. We thus repeated our analyses, but only included quiescent CD patients with a FCP <50 mcg/g, which has the lowest false negative rate for active inflammation in CD.^20^ Overall, our results were not changed by this lower threshold for FCP. Specifically, Shannon index was significantly decreased in qCD+symptoms compared with qCD-symptoms, IBS-D, and healthy controls (**Supplemental Figure 1A**, adjusted *P*=.01, *P*<.001, *P*<.0001, respectively). Similarly, microbial community structure was significantly different in qCD+symptoms compared with qCD-symptoms, IBS-D, and healthy controls (**Supplemental Figure 1B**, adjusted *P*=.001 for all comparisons).

### qCD+symptoms showed enrichment of oral taxa and depletion of butyrate/indole producers

We next examined which taxa were driving these microbial changes between groups. At the genus level, we identified 42, 52, and 43 taxa that were differentially abundant in qCD+symptoms relative to qCD-symptoms, HC, and IBS-D, respectively, while only one taxon was differentially abundant between qCD+symptoms and aCD (q<.05, **Supplemental Figure 2**).

At the species level and focusing specifically on qCD+symptoms vs. qCD-symptoms, we identified 63 species that were differentially abundant (q<.05, **Supplementary Table 1**). Of the species that demonstrated the greatest enrichment (log-fold change>2) in qCD+symptoms compared with qCD-symptoms, there were a striking number of species that are normal inhabitants of the oral microbiome, including *Klebsiella pneumoniae* and *Fusobacterium nucleatum* (**Figure 2A**). Conversely, the most significantly depleted species (log-fold change<-2) in qCD+symptoms compared with qCD-symptoms included several important butyrate and indole producers, including *Eubacterium rectale* and *Faecalibacterium prausnitzii*.

**Figure 2.**
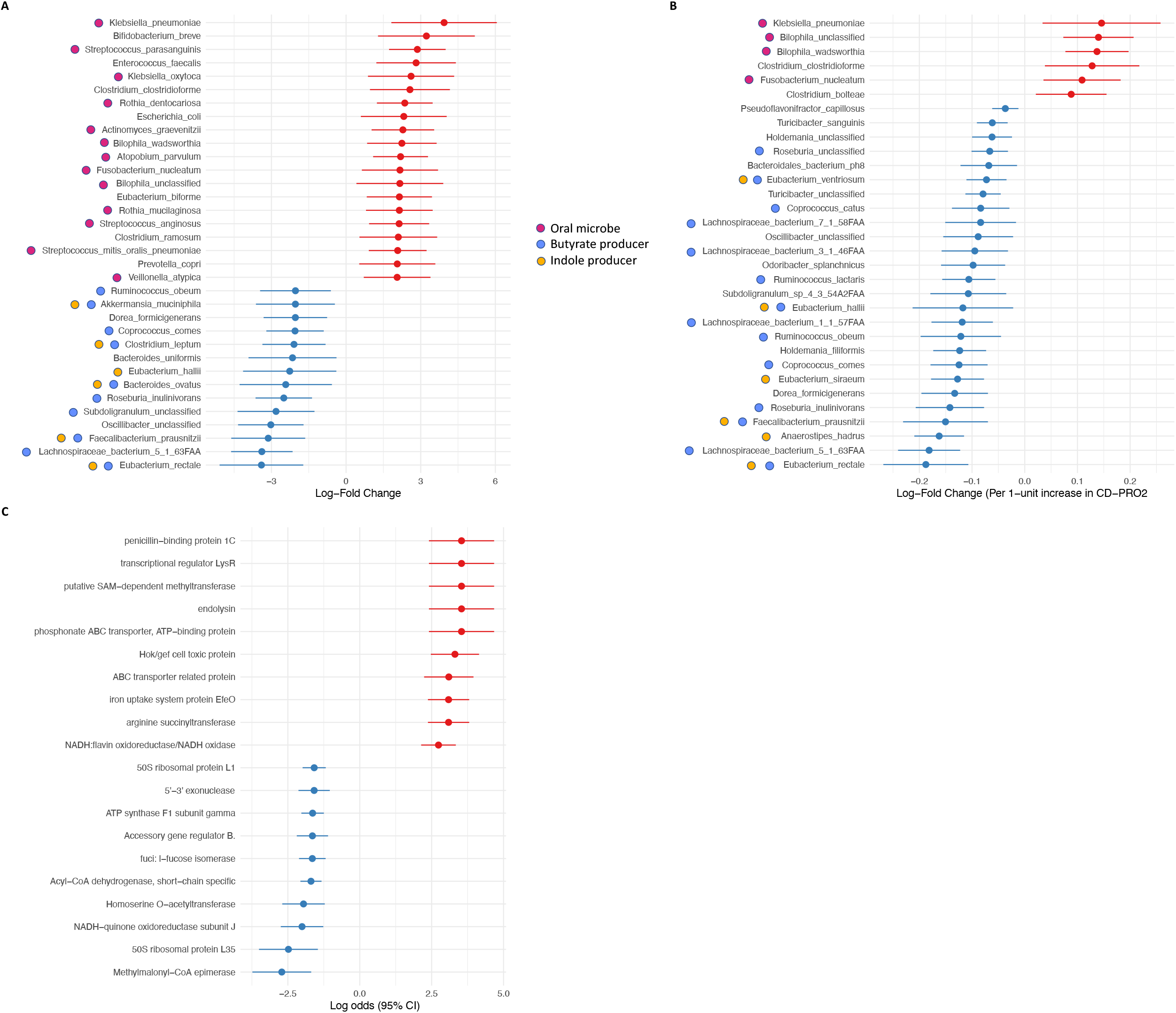
qCD+symptoms were enriched with normal inhabitants of the oral microbiome but depleted with important butyrate and indole producers. (**A**) The most differentially abundant species (q<.05, absolute log-fold change, lfc>2) in qCD+symptoms are shown. Species shown in r*ed* were enriched while those in *blue* were depleted in qCD+symptoms relative to qCD-symptoms. (**B**) Log-fold changes at the species level (q<.05) associated with CD-PRO2 score as a continuous outcome. (**C**) The most dysregulated microbial gene pathways (q<.05) by KEGG Orthology, including enriched (*red*) and depleted (*blue*) pathways, in qCD+symptoms compared with qCD-symptoms.

We also determined whether CD-PRO2 score was associated with microbial species by ANCOM-BC. Although our outcome here was modeled differently (i.e. as a continuous rather than a categorical outcome), we still found similar species were differentially abundant. Specifically, as CD-PRO2 score increases (indicating worse quality of life scores), we again found enrichment of species that are normal inhabitants of the oral microbiome as well as depletion of species important in butyrate and indole production (**Figure 2B**).

### Functional metagenome-related alterations in qCD+symptoms

We next identified potential functional changes in metagenomes using the KEGG Orthology (KO) database. There were significant differences in 152 KO groups between qCD+symptoms and qCD-symptoms (q<.05), including 91 enriched and 61 depleted (**Supplementary Table 2**). Pathways related to cysteine and methionine metabolism (SAM-dependent methyltransferase, homoserine O-acetyltransferase); ATP-binding cassette (ABC) transporters (phosphonate ABC transporter, ABC transporter related protein); fatty acid oxidation (methylmalonyl-CoA epimerase, acyl-CoA dehydrogenase); and iron transport and redox reactions (iron uptake system protein EfeO, NADH:flavin oxidoreductase/NADH oxidase, NADH-quinone oxidoreductase) were among the most dysregulated KO groups in qCD+symptoms compared to qCD-symptoms (**Figure 2C**).

### Bacterial indole pathways were altered in qCD+symptoms

We next focused on tryptophan pathways in qCD+symptoms. Tryptophan is catabolized by three major pathways, including kynurenine (Kyn), serotonin (5-HT), and indole (**Figure 3A**). While Trp metabolism to Kyn and 5-HT are mediated by both host and microbial enzymes, microbial enzymes are solely responsible for metabolism of tryptophan to indole derivatives.^13^ Thus, we concentrated our analyses on the bacterial *tnaA* gene encoding the enzyme tryptophanase, which catalyzes the first step in the conversion of Trp to indole. While bacterial *tnaA* alleles were ubiquitous in all groups, we found that CD had significantly fewer *tnaA* alleles compared with HC and IBS-D (adjusted *P*<.0001 for both; **Figure 3B**). Focusing on CD subgroups, qCD+symptoms had significantly fewer *tnaA* alleles compared with qCD-symptoms (adjusted *P*=.02) but not compared with aCD (adjusted *P*=.19).

**Figure 3.**
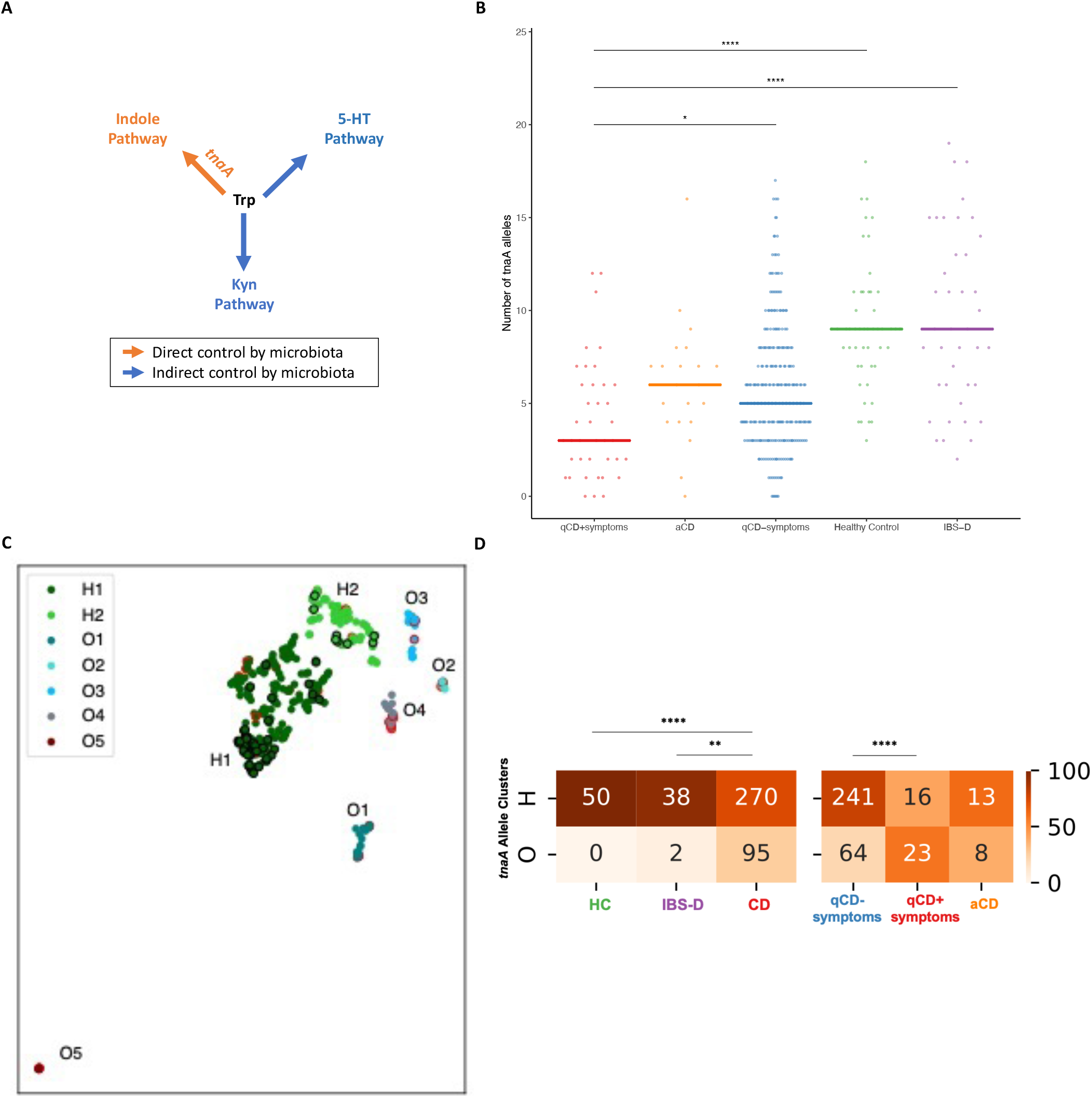
qCD+symptoms show reduced and significant allelic variation in t*naA* microbial genes. (**A**) Tryptophan (Trp) is catabolized via kynurenine (Kyn), serotonin (5-HT), and indole pathways. While Kyn and 5-HT pathways are mediated by both host and microbial enzymes, indole pathways are solely mediated by microbial enzymes, including tryptophanase (t*naA*). (**B**) The number of t*naA* alleles were significantly reduced in CD compared with HC and IBS-D. Focusing on CD subgroups, qCD+symptoms had significantly fewer *tnaA* alleles compared with qCD-symptoms but not for aCD. (**C**) Ordination of *tnaA* alleles by Jaccard distance using UMAP demonstrated 7 distinct clusters. Healthy controls (*black* outline) and IBS-D patients almost universally clustered into H1 and H2 (Healthy clusters) while CD patients, particularly qCD+symptoms (*red* outline), showed a greater distribution among clusters (Other O1-O5). (**D**) The proportion of CD patients clustering into the healthy clusters (H) was significantly different compared with HCs and IBS-D patients. These differences in clustering were mainly driven by qCD+symptoms compared with qCD-symptoms and aCD. Numbers represent the number of subjects from each group within each cluster while shading represents proportion of patients within each cluster. ** adjusted *P*<.005; **** adjusted *P*<.0001.

We then ordinated on Jaccard distance of *tnaA* alleles using UMAP and found that HC and IBS-D patients almost universally clustered into two clusters (Healthy H1, H2 clusters) (**Figure 3C**). There were significantly decreased number of CD patients clustering into H clusters (74.0%) compared with HCs (100%; adjusted *P*<.0001) and IBS-D patients (95.0%; adjusted *P*=.003, **Figure 3D**). Focusing on CD subgroups, we found that the proportion of qCD+symptoms clustering into H clusters was significantly lower compared with qCD-symptoms (41.0% vs. 79.0%, adjusted *P*<.0001, **Figure 3D**) but not compared to aCD (61.9%; adjusted *P*=.20).

### Microbiome predicted qCD+symptoms

To evaluate whether differences in microbial composition could be utilized to predict qCD+symptoms vs. qCD-symptoms, a random forest model was trained on microbial species abundance. The prediction of the model showed good accuracy by 5-fold cross-validation (AUC 0.73) as well as when validated using the independent test set (AUC 0.72) (**Figure 4A**). Interestingly, many of the species that were important to the model by variable importance scores (**Figure 4B**) were also found to be differentially abundant by ANCOM-BC (**Figure 2B**).

**Figure 4.**
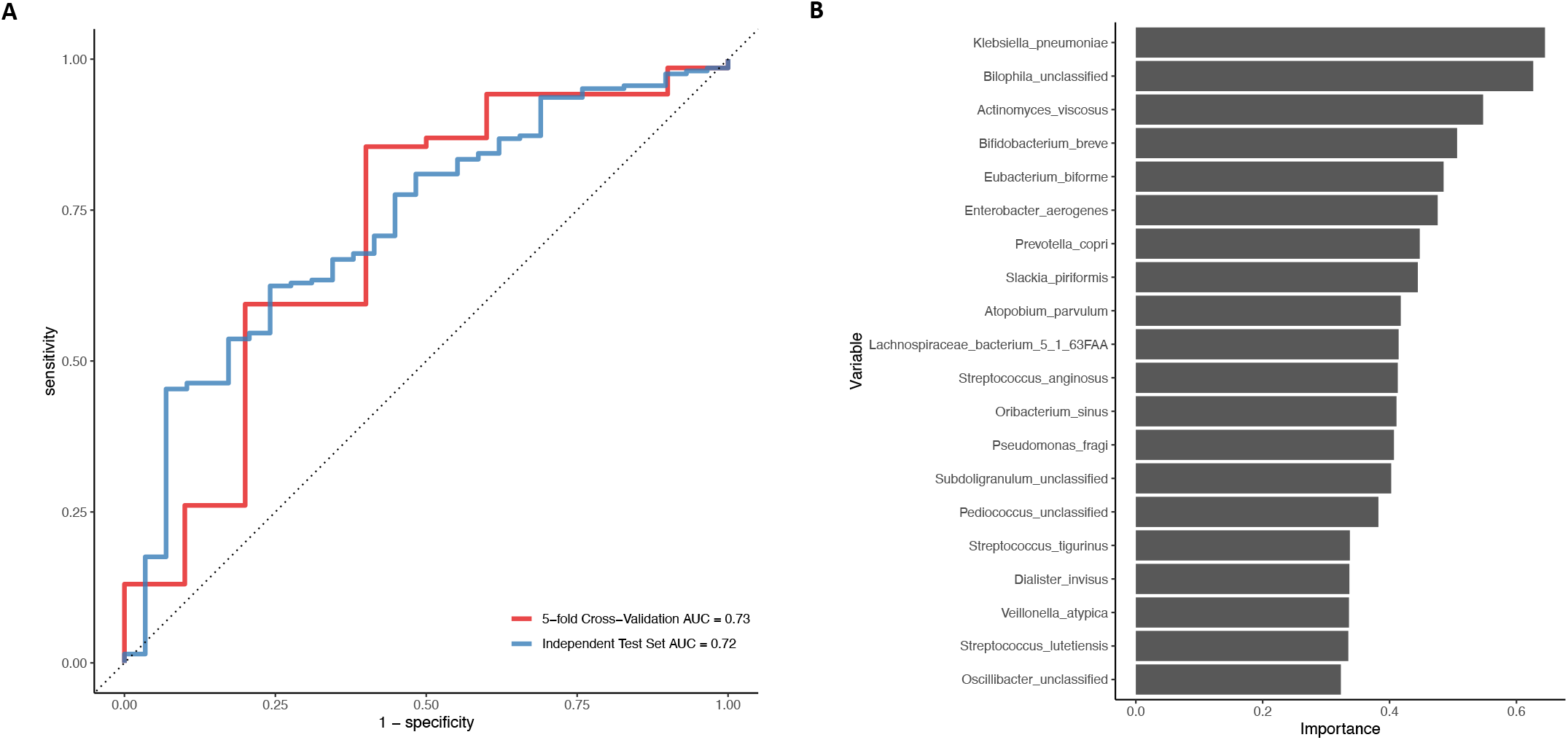
Microbial species accurately predicted quiescent CD with and without symptoms. (**A**) After randomly splitting the data into training (75%) and test (25%) set, a random forest model with 5-fold cross-validation was trained on species abundance. The accuracy of the model was evaluated by the area under the receiver operator characteristic curve (ROC). The model showed good accuracy for both the training data (AUC=0.73, *red line*) as well as when validated on the independent test set (AUC=0.72, *blue line*). (**B**) The top twenty species by variable importance scores are illustrated for the random forest model.

## Discussion

Although highly prevalent, the mechanisms underpinning persistent symptoms in quiescent CD are poorly understood. In this multi-center study, we demonstrated that patients with qCD+symptoms showed reduced microbial diversity, changes in microbial community structure and composition, and microbial function relative to qCD-symptoms, IBS-D, and HC. In contrast, the overall microbial community in qCD+symptoms was similar to aCD. Furthermore, qCD+symptoms demonstrated a striking enrichment of species normally found in the oral microbiome as well as depletion of important butyrate and indole producers. Additionally, microbial indole pathways were disrupted in qCD+symptoms, including depletion of microbial *tnaA* genes and evidence of *tnaA* allelic variation, compared with qCD-symptoms. Finally, we showed that microbial species were able to predict presence of qCD+symptoms with reasonable accuracy.

A novel finding in our study is that although qCD+symptoms share similar microbial diversity, community structure, and membership with aCD, there were significant differences in these microbial indices compared with qCD-symptoms. Indeed, while 63 taxa were differentially abundant between qCD+symptoms vs. qCD-symptoms, only 1 taxon was differentially abundant between qCD+symptoms and aCD. If we consider variation in the microbial community as a marker of host well-being that spans the continuum from health to disease, these findings suggest that the microbiome in qCD+symptoms is shifted towards an inflammatory phenotype while the community in qCD-symptoms may have reconstituted more towards healthy controls.

While prior studies have consistently demonstrated intestinal epithelial barrier dysfunction in qCD+symptoms,^7,8^ the mechanisms are largely unknown.^9^ We identified depletion of multiple bacterial species important in butyrate and indole production in qCD+symptoms, including *Akkermansia muciniphila, Faecalibacterium prausnitzii*, and *Roseburia, Lachnospiraceae*, and *Ruminococcus* spp.^13,21^ As butyrate and indole metabolites are important regulators of epithelial barrier integrity,^13,21^ these results suggest that altered gut microbiota may mediate epithelial barrier disruption in qCD+symptoms.

In line with these findings, we identified significant disruption in microbial-tryptophan pathways in qCD+symptoms compared with qCD-symptoms. As indole production is solely mediated by microbial enzymes, we focused our analysis on bacterial *tnaA* genes, which mediate the conversion of tryptophan to indole.^13^ We demonstrated that qCD+symptoms had significant depletion of *tnaA* alleles compared with qCD-symptoms. Furthermore, based on clustering of *tnaA* alleles, qCD+symptoms clustered distinctly and separately from qCD-symptoms. While most patients with qCD-symptoms clustered with HC and IBS patients, qCD+symptoms showed significantly higher membership outside of this healthy cluster. This allelic variation in bacterial *tnaA* genes suggest significant variability in the microbial community’s ability to compete for tryptophan and produce indole metabolites, which may contribute to intestinal barrier dysfunction in qCD+symptoms.

A novel finding in our study is the enrichment of bacterial species that are normal inhabitants of the oral microbiome. Normally, the oral microbiome is distinct from the gut microbiome in healthy adults.^22^ However, accumulating evidence supports the presence of an oral-gut axis with gut colonization of oral bacteria in CD.^23^ A recent study demonstrated that the oral and gut microbiome were more similar in CD patients, most of whom were in clinical remission, compared with healthy controls.^24^ Additionally, the presence of periodontitis increased future risk for flare in CD symptoms as measured by the short Crohn’s Disease Activity Index (sCDAI). As the sCDAI score is only weakly correlated with inflammation,^25^ this study may also have been demonstrating the relationship between oral-gut translocation of microbes and persistent symptoms in quiescent CD. Although the functional consequences of oral-gut translocation in qCD+symptoms are unknown, cysteine and methionine metabolism, ATP transport, and redox reactions were among the most highly dysregulated pathways in qCD+symptoms. Cysteine and methionine are two primary sulfur-containing amino acids, while many oral microbes are known to be producers of hydrogen sulfide (H_2_S). As H_2_S is an important regulator of mitochondrial function, oxidative stress, and intestinal permeability,^26,27^ these data are suggestive of a novel pathway in qCD+symptoms.

There are several strengths of our work. First, we utilized data from the multicenter, prospective SPARC IBD cohort. Given the variability of the microbiome in CD,^28^ this strategy of sampling from a geographically, racially, and ethnically diverse cohort across the US helps to generalize our findings. Furthermore, the highly standardized data and biosample collection methods are also strengths, which reduces bias and improves reproducibility.^14^ In addition, the inclusion of data from active CD and non-IBD patients allowed us to contextualize how the microbiome varies from active inflammation to health. Finally, while common metagenomic approaches are limited by the robustness of reference databases, our bioinformatics approach focused on microbial protein-coding genes that are assembled de novo and thus do not have these same constraints. Furthermore, this approach allowed us to evaluate our *a priori* hypothesis on bacterial *tnaA* genes and infer the metagenomic potential of the microbial community.

However, there are limitations to our work as well. First, only a subset of patients had endoscopic evidence of quiescent disease, and thus we cannot completely exclude the possibility that our results were influenced by inflammation. However, our results remained robust even when we restricted our analysis to patients with FCP <50 mcg/g, which has the lowest false negative rate in detecting inflammation in CD.^20^ These results suggest that the microbiome changes described here are likely independent from inflammation. Secondly, while these results support an association between microbial changes and persistent symptoms in quiescent CD, we cannot make any firm conclusions on causation here. Similarly, we do not have longitudinal data to suggest directionality between microbial changes and persistent symptoms. Finally, although diet is known to be a major influence on the microbiome, we did not have dietary information on CD patients.

In conclusion, we have identified quiescent CD patients with persistent symptoms have widespread changes in their intestinal microbiota similar to the microbial community found in active CD but quite distinct from quiescent CD without persistent symptoms. Specifically, quiescent CD patients with persistent symptoms had enrichment of bacterial species normally found in the oral microbiome as well as depletion of species important in butyrate and tryptophan metabolism. These proof-of-concept findings suggest an altered microbiome may mediate intestinal barrier dysfunction, which has been consistently demonstrated in prior studies. While our findings require validation, future work may determine whether microbiome-based therapies show promise as novel therapeutic targets for persistent symptoms in quiescent CD.

## Data Availability

All data produced in the present work are contained in the manuscript

---

|  | SPARC IBD Cohort |  |  | Non-IBD Cohort |  |
| --- | --- | --- | --- | --- | --- |
|  | Active CD<br>(n=21) | qCD+symptoms<br>(n=39) | qCD-symptoms<br>(n=274) | IBS-D<br>(n=40) | Healthy<br>controls<br>(n=50) |
| Age, mean (sd) | 40.0 (14.3) | 50.3 (14.2) | 43.2 (15.1) | 45.6 (18.3) | 46.9 (15.5) |
| Female gender, n (%) | 13 (61.9) | 28 (71.8) | 168 (61.3) | 29 (72.5) | 16 (32.0) |
| Race, n (%) |  |  |  |  |  |
| White | 15 (78.9) | 31 (91.2) | 225 (92.6) | 37 (92.5) | 37 (74.0) |
| Black | 3 (15.8) | 2 (5.9) | 12 (4.9) | 2 (5.0) | 2 (4.0) |
| Other | 1 (5.3) | 1 (2.9) | 6 (2.5) | 1 (2.5) | 11 (22.0) |
| Ethnicity, n (%) |  |  |  |  |  |
| Hispanic/Latino | 2 (10.5) | 0 (0.0) | 2 (0.8) | 0 (0.0) | 3 (6.0) |
| Non-Hispanic/Latino | 17 (89.5) | 32 (97.0) | 236 (97.1) | 40 (100.0) | 41 (82.0) |
| Other | 0 (0.0) | 1 (3.0) | 5 (2.1) | 0 (0.0) | 6 (12.0) |
| BMI (kg/m<sup>2</sup>), mean (sd) | 27.2 (6.1) | 32.1 (8.0) | 28.1 (6.5) | 28.5 (6.8) | 28.0 (4.4) |
| CD Location, n (%) |  |  |  |  |  |
| Large intestine | 2 (9.5) | 3 (7.7) | 40 (14.6) |  |  |
| Small and large intestine | 13 (61.9) | 23 (59.0) | 130 (47.4) |  |  |
| Small intestine | 3 (14.3) | 4 (10.3) | 56 (20.4) |  |  |
| Other | 3 (14.3) | 9 (23.1) | 48 (17.5) |  |  |
| Current CD Medication, n (%) |  |  |  |  |  |
| 5-ASA | 2 (9.5) | 2 (5.4) | 34 (12.5) |  |  |
| Steroids | 11 (52.4) | 8 (21.6) | 20 (7.3) |  |  |
| Immunomodulators | 5 (23.8) | 11 (29.7) | 88 (32.2) |  |  |
| Biologics | 15 (71.4) | 28 (75.7) | 189 (69.0) |  |  |
| None | 2 (9.5) | 5 (13.5) | 32 (11.7) |  |  |
| Prior IBD surgery, n (%) | 6 (35.3) | 18 (52.9) | 60 (26.3) |  |  |
| Total SES-CD score, mean (sd) | 9 (15.6) | 1.8 (2.0) | 2.3 (4.1) |  |  |
| Fecal calprotectin, mean (sd) | 1017.9 (639.1) | 49.7 (41.8) | 41.3 (41.8) |  |  |
| Serum CRP level, mean (sd) | 10.0 (15.5) | 2.6 (3.5) | 1.3 (2.3) |  |  |
| History of depression, n (%) | 7 (46.7) | 11 (33.3) | 60 (27.5) |  |  |
| History of anxiety, n (%) | 6 (40.0) | 15 (45.5) | 58 (26.9) |  |  |

BMI, body mass index (kg/m<sup>2</sup>); CD, Crohn's Disease; CRP, C-reactive protein; IBD, inflammatory bowel disease; SES-CD, simple endoscopic score for Crohn's disease

## References

1. Olivera P, Danese S, Peyrin-Biroulet L. Next generation of small molecules in inflammatory bowel disease. Gut 2017;66:199–209.

2. Halpin SJ, Ford AC. Prevalence of symptoms meeting criteria for irritable bowel syndrome in inflammatory bowel disease: systematic review and meta-analysis. Am J Gastroenterol 2012;107:1474–1482.

3. Gracie DJ, Williams CJM, Sood R, et al. Negative Effects on Psychological Health and Quality of Life of Genuine Irritable Bowel Syndrome–type Symptoms in Patients With Inflammatory Bowel Disease. Clinical Gastroenterology and Hepatology 2017;15:376–384.e5.

4. Anderson A, Click B, Ramos-Rivers C, et al. The Association Between Sustained Poor Quality of Life and Future Opioid Use in Inflammatory Bowel Disease. Inflamm Bowel Dis 2018;24:1380–1388.

5. Lichtenstein GR, Feagan BG, Cohen RD, et al. Serious infection and mortality in patients with Crohn’s disease: more than 5 years of follow-up in the TREATTM registry. Am J Gastroenterol 2012;107:1409–1422.

6. Limsrivilai J, Stidham RW, Govani SM, et al. Factors That Predict High Health Care Utilization and Costs for Patients With Inflammatory Bowel Diseases. Clin Gastroenterol Hepatol 2017;15:385–392.e2.

7. Chang J, Leong RW, Wasinger VC, et al. Impaired Intestinal Permeability Contributes to Ongoing Bowel Symptoms in Patients With Inflammatory Bowel Disease and Mucosal Healing. Gastroenterology 2017;153:723–731.e1.

8. Vivinus-Nébot M, Frin-Mathy G, Bzioueche H, et al. Functional bowel symptoms in quiescent inflammatory bowel diseases: role of epithelial barrier disruption and low-grade inflammation. Gut 2014;63:744–752.

9. Henriksen M, Høivik ML, Jelsness-Jørgensen L-P, et al. Irritable Bowel-like Symptoms in Ulcerative Colitis are as Common in Patients in Deep Remission as in Inflammation: Results From a Population-based Study [the IBSEN Study]. Journal of Crohn’s and Colitis 2018;12:389–393.

10. Boland K, Bedrani L, Turpin W, et al. Persistent Diarrhea in Patients With Crohn’s Disease After Mucosal Healing Is Associated With Lower Diversity of the Intestinal Microbiome and Increased Dysbiosis. Clin Gastroenterol Hepatol 2021;19:296–304.e3.

11. Humbel F, Rieder JH, Franc Y, et al. Association of Alterations in Intestinal Microbiota With Impaired Psychological Function in Patients With Inflammatory Bowel Diseases in Remission. Clin Gastroenterol Hepatol 2020;18:2019–2029.e11.

12. Borren NZ, Plichta D, Joshi AD, et al. Alterations in Fecal Microbiomes and Serum Metabolomes of Fatigued Patients With Quiescent Inflammatory Bowel Diseases. Clin Gastroenterol Hepatol 2021;19:519–527.e5.

13. Roager HM, Licht TR. Microbial tryptophan catabolites in health and disease. Nature Communications 2018;9:3294.

14. Raffals LE, Saha S, Bewtra M, et al. The Development and Initial Findings of A Study of a Prospective Adult Research Cohort with Inflammatory Bowel Disease (SPARC IBD). Inflamm Bowel Dis 2022;28:192–199.

15. Turner D, Ricciuto A, Lewis A, et al. STRIDE-II: An Update on the Selecting Therapeutic Targets in Inflammatory Bowel Disease (STRIDE) Initiative of the International Organization for the Study of IBD (IOIBD): Determining Therapeutic Goals for Treat-to-Target strategies in IBD. Gastroenterology 2021;160:1570–1583.

16. Khanna R, Zou G, D’Haens G, et al. A retrospective analysis: the development of patient reported outcome measures for the assessment of Crohn’s disease activity. Alimentary Pharmacology & Therapeutics 2015;41:77–86.

17. Minot SS, Barry KC, Kasman C, et al. geneshot: gene-level metagenomics identifies genome islands associated with immunotherapy response. Genome Biol 2021;22:135.

18. Lin H, Peddada SD. Analysis of compositions of microbiomes with bias correction. Nat Commun 2020;11:3514.

19. Beghini F, McIver LJ, Blanco-Míguez A, et al. Integrating taxonomic, functional, and strain-level profiling of diverse microbial communities with bioBakery 3. eLife 10:e65088.

20. Kopylov U, Yung DE, Engel T, et al. Fecal calprotectin for the prediction of small-bowel Crohn’s disease by capsule endoscopy: a systematic review and meta-analysis. Eur J Gastroenterol Hepatol 2016;28:1137–1144.

21. Agus A, Planchais J, Sokol H. Gut Microbiota Regulation of Tryptophan Metabolism in Health and Disease. Cell Host & Microbe 2018;23:716–724.

22. Rashidi A, Ebadi M, Weisdorf DJ, et al. No evidence for colonization of oral bacteria in the distal gut in healthy adults. Proc Natl Acad Sci USA 2021;118:e2114152118.

23. Gevers D, Kugathasan S, Denson LA, et al. The Treatment-Naive Microbiome in New-Onset Crohn’s Disease. Cell Host & Microbe 2014;15:382–392.

24. Imai J, Ichikawa H, Kitamoto S, et al. A potential pathogenic association between periodontal disease and Crohn’s disease. JCI Insight 2021;6:e148543.

25. Jones J, Loftus EV, Panaccione R, et al. Relationships between disease activity and serum and fecal biomarkers in patients with Crohn’s disease. Clin Gastroenterol Hepatol 2008;6:1218–1224.

26. Blachier F, Beaumont M, Kim E. Cysteine-derived hydrogen sulfide and gut health: a matter of endogenous or bacterial origin. Curr Opin Clin Nutr Metab Care 2019;22:68–75.

27. Murphy B, Bhattacharya R, Mukherjee P. Hydrogen sulfide signaling in mitochondria and disease. FASEB J 2019;33:13098–13125.

28. Clooney AG, Eckenberger J, Laserna-Mendieta E, et al. Ranking microbiome variance in inflammatory bowel disease: a large longitudinal intercontinental study. Gut June 2020.

