## Supplementary figures and images for "The Fecal Microbiome in Quiescent Crohn’s Disease with Persistent Gastrointestinal Symptoms Show Enrichment of Oral Microbes But Depletion of Butyrate and Indole Producers"

### Supplemental Figure 1

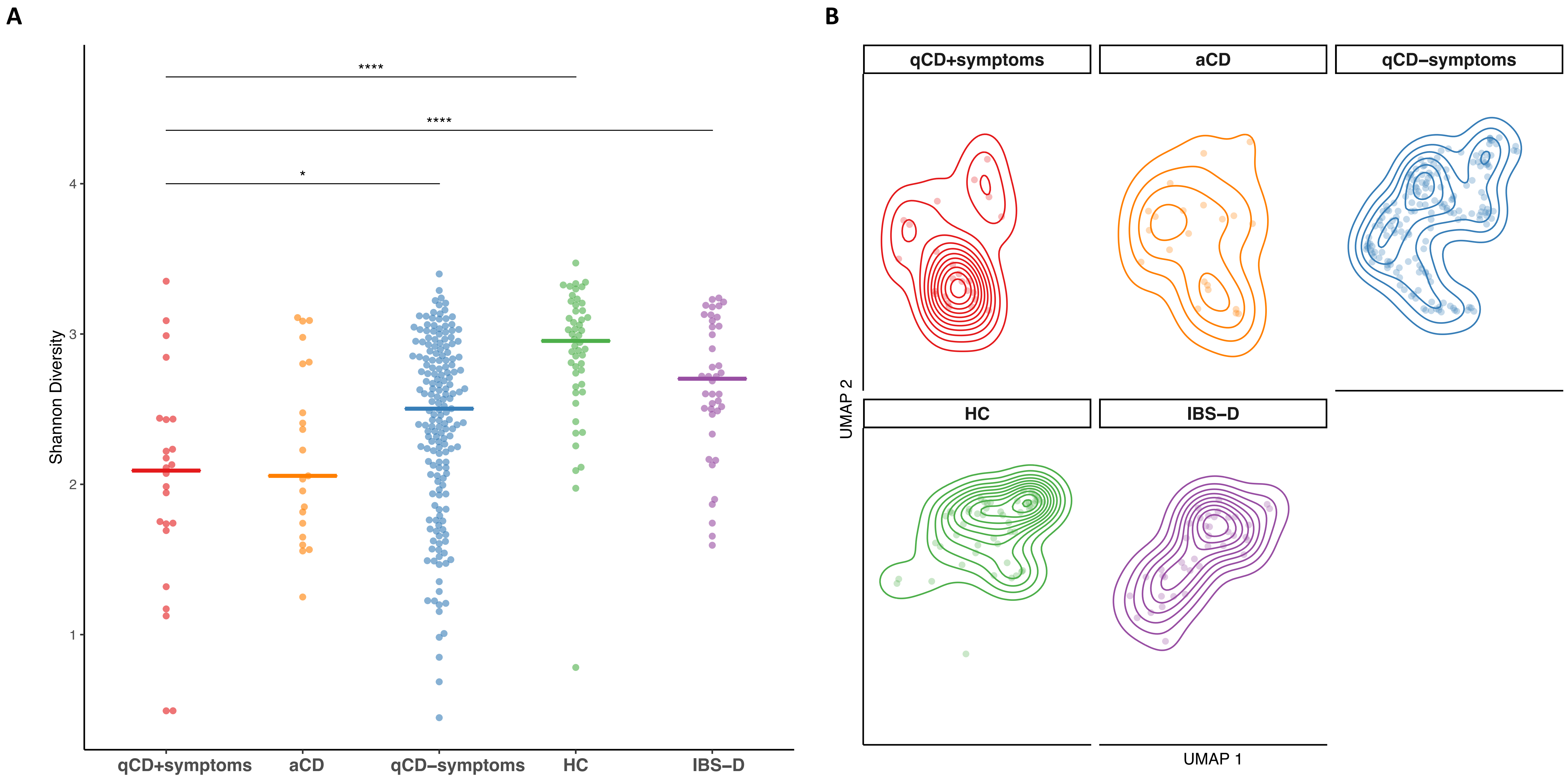

### Supplemental Figure 2

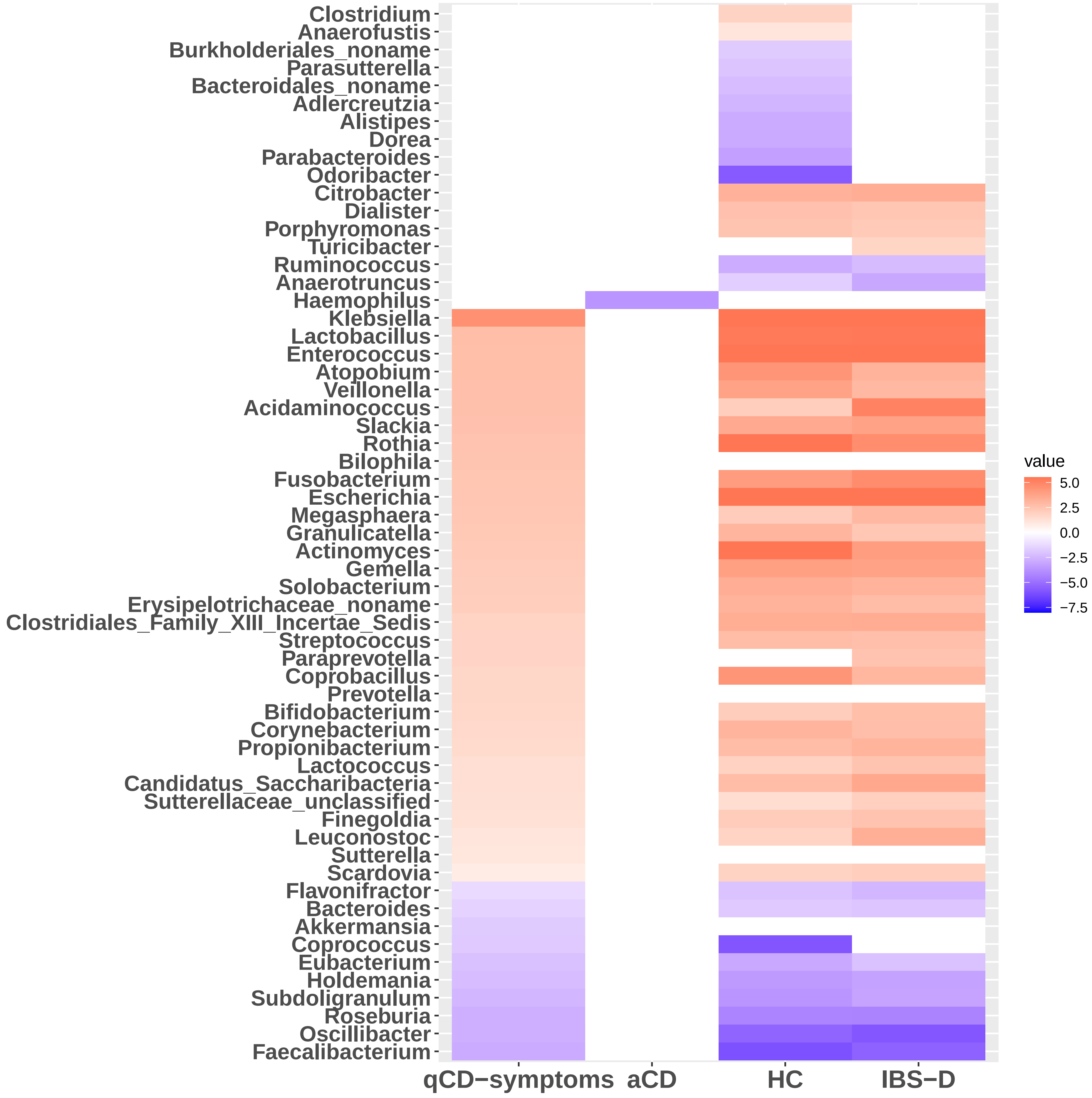
